# Integrating epidemiological and genetic data with different sampling intensities into a dynamic model of respiratory syncytial virus transmission

**DOI:** 10.1101/2020.03.08.20030742

**Authors:** Ivy K. Kombe, Charles N. Agoti, Patrick K. Munywoki, D. James Nokes, Graham F. Medley

## Abstract

**Background:** Respiratory syncytial virus (RSV) is responsible for a significant burden of acute respiratory illness in children under 5 years old. Prior to rolling out any vaccination program, identification of the source of infant infections could further guide vaccination strategies.

**Methods:** We extended a dynamic model calibrated at the individual host level initially fit to social-temporal data on shedding patterns to include whole genome sequencing data available at a lower sampling intensity.

**Results:** In this study population of 493 individuals with 55 infants under the age of 1 year distributed across 47 households, we found that 52% of RSV-B and 60% of RSV-A cases arose from infection within the household. Forty-five percent of infant infections appeared to occur in the household, of which 68% were a result of transmission from a child aged between 2 and 13 years living in the same household as the infant.

**Conclusion:** These results further highlight the importance of pre-school and school-aged children in RSV transmission, particularly the role they play in directly infecting the household infant. These age groups are a potential RSV vaccination target group.

## Introduction

In 2015 the estimated RSV acute lower respiratory illness (ALRI) burden in children less than 5 years old was 33.1 million cases resulting in 118,200 (94,600-149,400) deaths. Over 90% of the estimated RSV burden was in developing countries [1]. A recent study across sites in 7 low-income and low-middle-income countries looking into the aetiology of severe and very severe pneumonia found that RSV is the single pathogen with the largest attributable fraction [2]. Infants below 6 months of age experience the most severe disease [3]. Increasingly, RSV is also being identified as a disease causing pathogen in the elderly [4]. There are currently over fifty candidate vaccines against RSV at different stages of development with the most advanced being a maternal vaccine [5–7].

RSV disease occurs in a seasonal pattern with most populations experiencing annual cycles [8–12]. Virus isolates can be classified into two antigenically and genetically distinct groups (RSV-A and RSV-B) and consecutive seasons are not only characterized by a change in the dominant group, but also changes to the genotype composition [12,13]. Though several studies have predicted a maternal vaccination would be effective [14–16] by extending the duration of protection by passive immunity early in life, the vaccination of older children has also been theorized as an effective alternative or complementary strategy by producing a heard immunity effect [17–20]. Elder and, particularly, school going children have been shown in previous work to be associated with increased risk of infant (sibling) infection [21– 24] - though no direct infection link between the older siblings and the infant was confirmed - and have been identified as drivers of the initial epidemic phase [25]. Identifying the role of different age and social groups in RSV transmission networks may provide further evidence for optimal vaccine target groups.

Previously, using data from a cohort study that followed household members for 6 months, we have attempted to identify the source of infant infection in the household. In a descriptive analysis of the social-temporal data, school-going siblings were frequently (73%) identified as index cases in household outbreaks where an infant was infected [24]. In a phylogenetic analysis of whole genome sequence (WGS) data from of a subset of the household data the household source of infant infection was definitively identified for just 4 of the 23 infant cases in the subset data, while 9 others were identified as index cases in household outbreaks [26]. An attempt to use shared minor variants obtained from deep sequencing failed to add further resolution to the transmission chains [27]. In a modelling study using the social-temporal data, it was found that about half of all cases occurred in the household [28]. Independently, these studies were unable to clearly determine who infected the infants with RSV and how infection spread once introduced in the household.

In this article, we extend the previous modelling study to integrate social-temporal and WGS data to identify generalizable characteristics of RSV transmission chains at the household level. In doing so, we will identify if data integration, and hence increased pathogen resolution, increases the precision with which model parameters are estimated or changes the estimates such that different transmission dynamics are inferred. To our knowledge, this is the first attempt at combining these two data types in a single modelling framework for RSV. There are several approaches to integrating genetic data with other data types [29–31], the choice of which is dependent on the data available and the aims of the study [32]. Similar to the approach used by Didelot et al, [33] we used a two-step approach of first making inference from the genetic data and then incorporating this into the dynamic transmission model of RSV.

## Methods

### Data

During a seasonal RSV outbreak beginning late 2009, members of 47 households in a rural coastal Kenya were followed up for a period of 6 months with an aim of recording the incidence of RSV and inferring who infects the infant [24]. A household in this study was defined as comprising of people who share food from the same kitchen. Households were recruited on the basis of having an infant born after the previous RSV epidemic who had at least 1 elder sibling <13 years old. Members of the household had nasopharyngeal swab (NPS) samples and clinical data collected every 3-4 days. The samples were tested for RSV using an in-house real-time multiplexed polymerase chain reaction (PCR) assay [34]. A sample was considered RSV positive if the PCR cycle threshold value was >0 and ≤35. An RSV infection episode was defined as a period within which an individual provided positive samples for the same RSV group that were no more than 14 days apart. A shedding episode was referred to as symptomatic if within the window of virus shedding, there is at least one day where symptoms were recorded. The symptoms of interest are those of an acute respiratory illness (ARI), which are: cough, or nasal discharge/blockage, or difficulty breathing. There were 16928 samples collected, of which 205 were positive for RSV-A and 306 for RSV-B. This translated to 97 RSV-A episodes (88 infected individuals and 25 infected households) and 125 RSV-B episodes (113 infected individuals and 34 infected households).

Whole genome sequences (WGS) were obtained for 103 (41.2%) of the RSV-A samples and 88 (28.8%) of the RSV-B using the Illumina MiSeq platform [26]. The sequences were distributed across 54 (55.6%) episodes, 50 (56.8%) individuals and 9 (36%) households for RSV-A, and 54 (43.2%) episodes, 53 (45.9%) individuals and 15 (44.1%) households for RSV-B. During phylogenetic analysis [26], genetic clades and subclades were established based on a combination of criteria: nucleotide distance cut-off, clustering patterns on the global RSV phylogeny and the inferred date of sequence divergence. We did not make a distinction between clades and subclades, resulting in 5 RSV-A and 7 RSV-B clusters.

The model required daily infection data where a viral shedding episode can be identified by RSV group and by genetic cluster within each group. However, given the sampling interval and incomplete sequencing, we had to make assumptions to fill in the days of missing data. Imputation of cluster shedding durations was done for episodes that had at least one sequence, and for episodes with no sequences yet were part of a household outbreak with at least one sequence. The cluster ID for any episodes left unassigned at this stage were inferred along with the model parameters. Details of the data pre-processing can be found in supplementary appendix A1.

Informed written consent was obtained from all the study participants or their parent/guardian. The KEMRI-Scientific and Ethical Review Committee in Kenya provided ethical approval for the initial study and any analysis thereafter. The Observational/ Interventions Research Ethics Committee at the London School of Hygiene and Tropical Medicine provided further approval for this analysis.

### Transmission model

Similar to our previous work [28], we modelled RSV transmission at the individual host level in daily timesteps for the 6-month study period. The model was formulated to investigate the factors that determine infection onset, following from which, inference on the transmission chain was made.

Everyone was assumed to be susceptible to infection by RSV at the start of the outbreak, but the risk of infection was dependent on age. Once individuals were exposed to infection, they entered a latency period that ranged between 2 to 5 days after which they became infectious [35]. After the infectious period, individuals became susceptible to infection again, but with a modified risk, i.e. RSV conferred partial transient immunity that lasts as long as the outbreak is ongoing. This partial immunity is assumed to be different for heterologous group re-infection and homologous group re-infection. Individuals can get heterologous group co-infections, we explored if susceptibility to infection by RSV-A was modified if an individual was currently shedding RSV-B, and vice-versa.

The main assumptions about transmission were contained in the equation giving the cluster specific (index ***c***) per capita (index ***i***) rate of exposure to infection per unit time (denoted ***t***), also known as the infection hazard or force of infection, denoted *λ*_*i,c*_*(t)*. At its base:

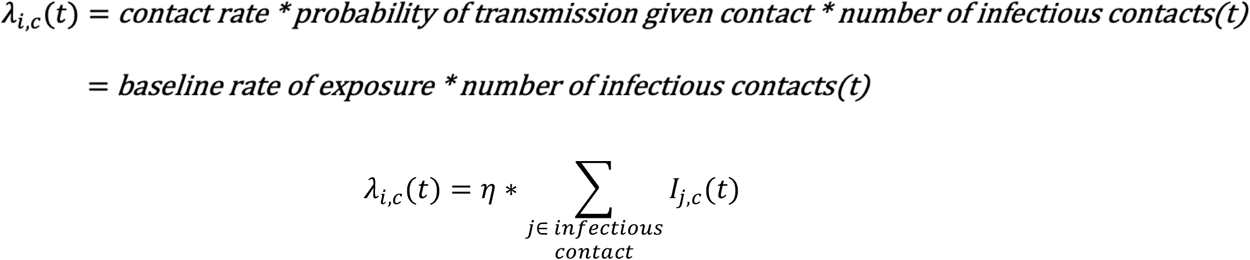

Where *η* is the baseline rate of exposure and *I*_*j,c*_*(t)* is an indicator variable of infectiousness of ***j*** at time ***t***.

In addition to the assumptions about RSV natural history, we extend this basic formulation to allow transmission from within the household or external sources, and to explore if factors such as household size, infectiousness (as determined by viral load and ARI symptoms) and age are determinants of exposure. We also explored if within-cluster genetic (nucleotide) distances could further be used to identify transmission events. The final model had 19 parameters. Further model details can be found in supplementary appendix A2.

We used Bayesian inference and Metropolis-Hasting Markov Chain Monte Carlo (MH-MCMC) to obtain estimates of the model parameters and augment missing cluster identities given the observed data. Further details can be found in supplementary appendix A3. All the computation was done using the *Julia* language (version 1.1) [36,37]. The code is publicly available at https://github.com/Ikadzo/HH_Transmission_Model.

Following the estimation of the posterior parameter distribution, we randomly selected a subset to determine infection sources for every case. Further details can be found in the supplementary appendix A4. A hundred parameter sets were sampled and the highest probability transmission source (HPTS) for each case established for each sample. From the distribution of 100 HPTS, the one with the highest frequency was selected as the source of transmission. This frequency becomes the weight assigned in the transmission network.

## Results

The data imputation process resulted in shedding episodes that ranged from 2 to 35 days for RSV A, and 3 to 45 days for RSV B. The cluster ids for 12 of 43 RSV A episodes and 19 of 71 RSV B episodes with no genetic information were imputed prior to model fitting, the rest were inferred along with model parameters. The shedding patterns after the data pre-processing are shown in Figure 1 and Figure 2. The study initially recruited 60 households but 13 were lost to follow-up, hence the numbering of the households goes beyond 47.

**Figure 1:**
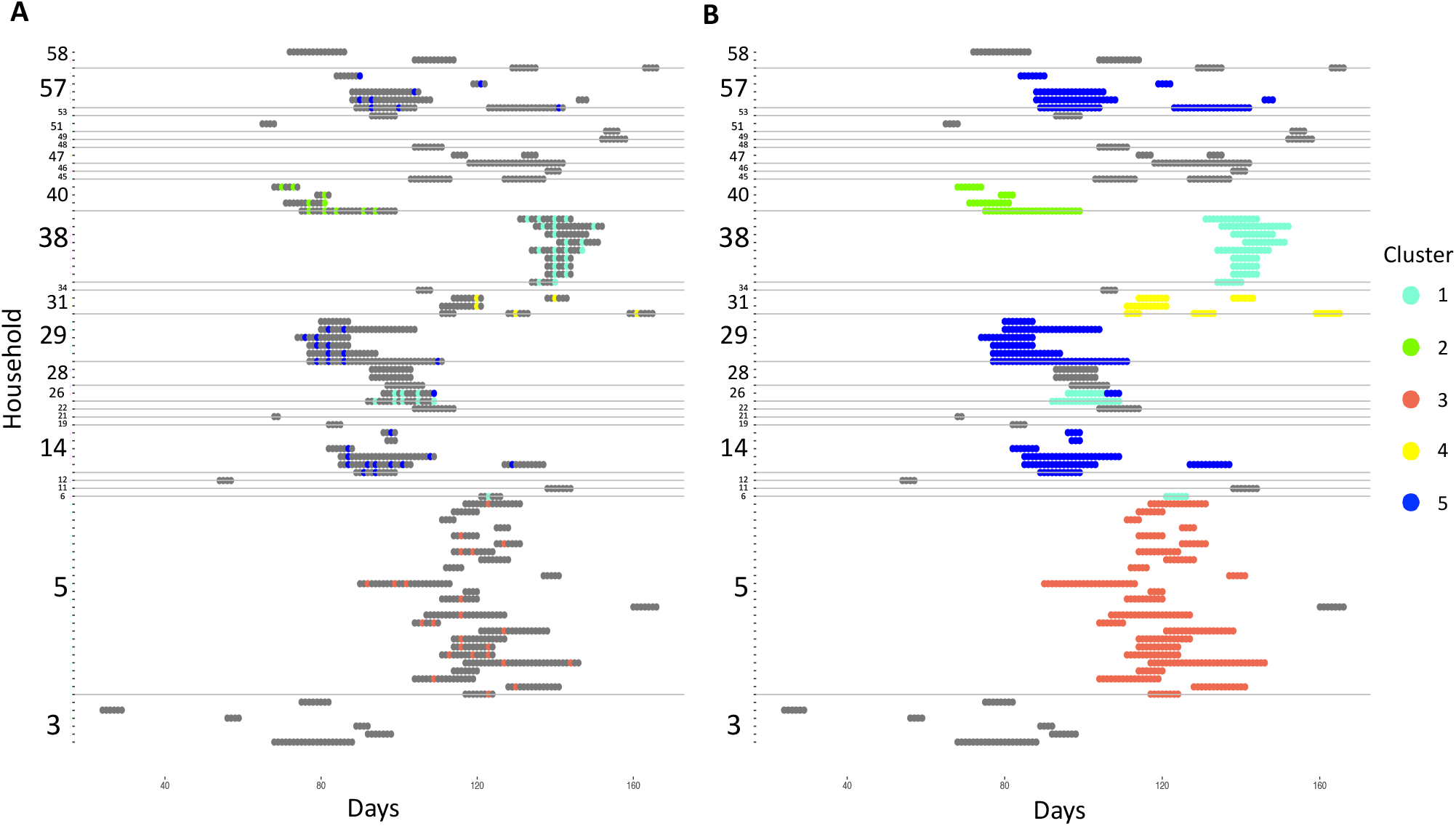
Distribution of available sequences across RSV-A infection episodes (A) and the results of the imputation of genetic information. **(B)**. The y-axis shows the household where each notch is a single individual, time in days is on the x-axis. The horizontal grey lines demarcate the households. The grey regions shows days of shedding whose cluster id is unknown.

**Figure 2:**
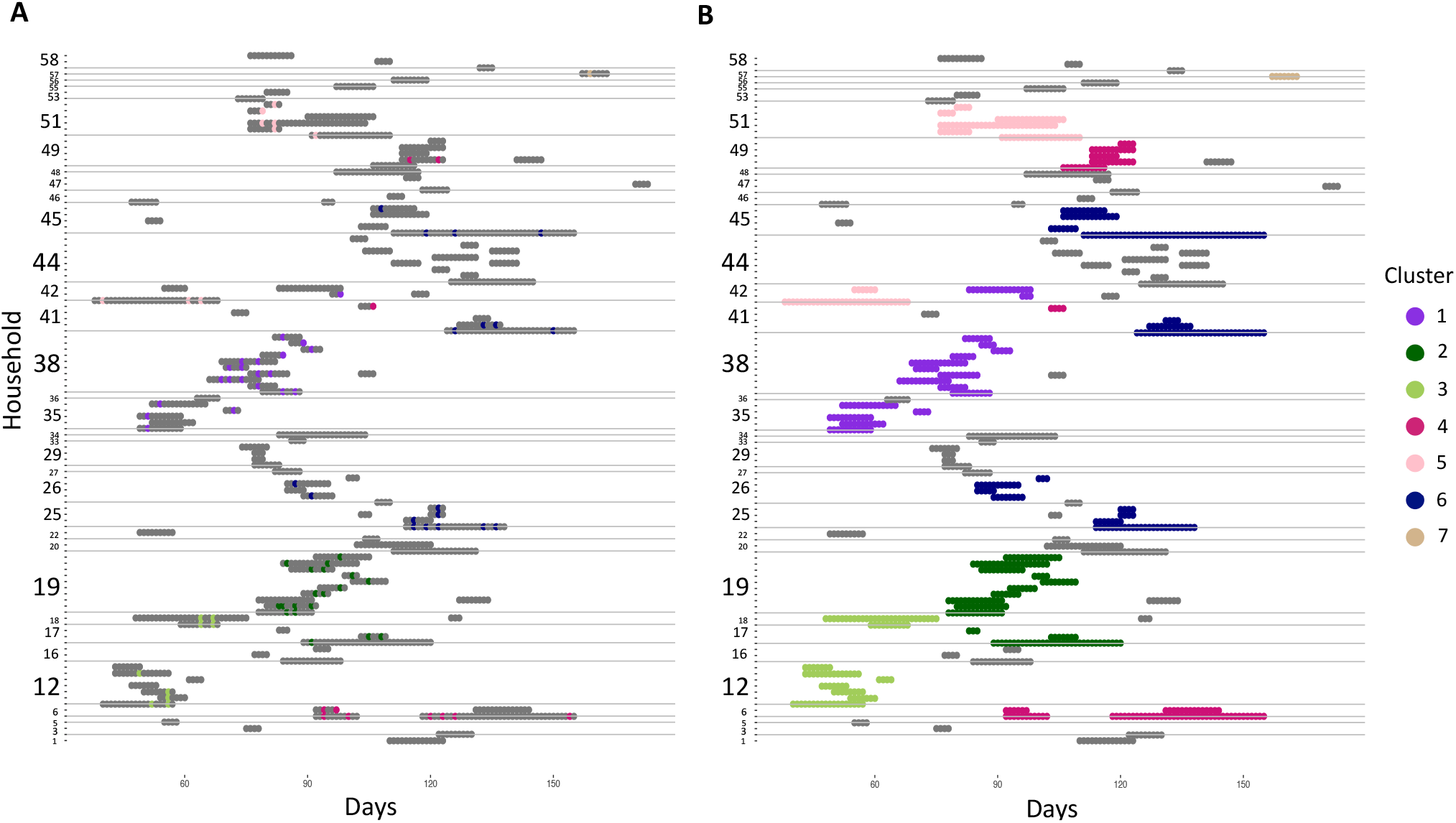
Distribution of available sequences across RSV-B infection episodes (A) and the results of the imputation of genetic information. **(B)**. The y-axis shows the household where each notch is a single individual, time in days is on the x-axis. The horizontal grey lines demarcate the households. The grey regions shows days of shedding whose cluster id is unknown.

### Model Inference

Parameter trace plots and results of convergence checks can be found in supplementary appendix A5. Based on the estimated parameters we observed an inverse relationship between age and susceptibility to infection, relative to infants (<1 year old), the percentage reduction in the rate of exposure for 1-4, 5-14 and ≥15 year olds was 22% (95% CrI:50%, - 61%), 73% (95% CrI:51%, 84%) and 84% (95% CrI:71%, 91%), respectively. RSV conferred partial immunity following infection, more so for homologous (57% (95% CrI:33%, 73%) reduced exposure) than heterologous (49% (95% CrI:10%, 74%) reduced exposure) group reinfections. Households of ≥ 8 individuals had a 54% (95% CrI:32%, 71%) reduction in pair-wise rate of exposure within the household relative to smaller households. Symptomatic cases were more infectious than asymptomatic cases, more so with a high viral load (relative infectiousness 4.4 (95% CrI:1 .8, 9.0)) than a low viral load (relative infectiousness: 2.1 (95% CrI: 1.2, 3.7)). Transmission between the study households is unlikely to have occurred, although the large credibility interval on the parameter suggests that there is limited information for this parameter. Estimates of the rate of exponential decrease in transmission probability with increasing genetic distance parameter imply that within cluster transmission was nearly 100% likely regardless of the pair-wise nucleotide distances. The effect of age in community exposure was unclear as the parameters were estimated with credible intervals including 1. Table A5.3 of parameter estimates can be found in supplementary appendix A5.

To validate the model, we simulated multiple epidemics which verified that key aspects of the epidemic were being reproduced by the simulations. Details of this can be found in supplementary appendix A6.

To assess the impact of increased resolution in pathogen identification on estimated parameters we compared the distributions of parameters estimated using RSV cases identified at the pathogen, group and cluster level. Figure 3 shows the density plots comparing these distributions, details of the model modifications to allow fitting of group level data are in supplementary appendix A7. This figure shows 17 of the 19 comparable parameters in the model with genetic clusters. For most of the parameters, the estimated distributions do not differ by resolution in pathogen identification. The parameters measuring the effect of viral load and symptoms on infectiousness are estimated with increased precision with increased pathogen resolution. The distribution of the within household transmission coefficients shift slightly towards higher values with increased resolution both for RSV-A and B while the community transmission coefficient for RSV-A has a slight shift towards lower values.

**Figure 3:**
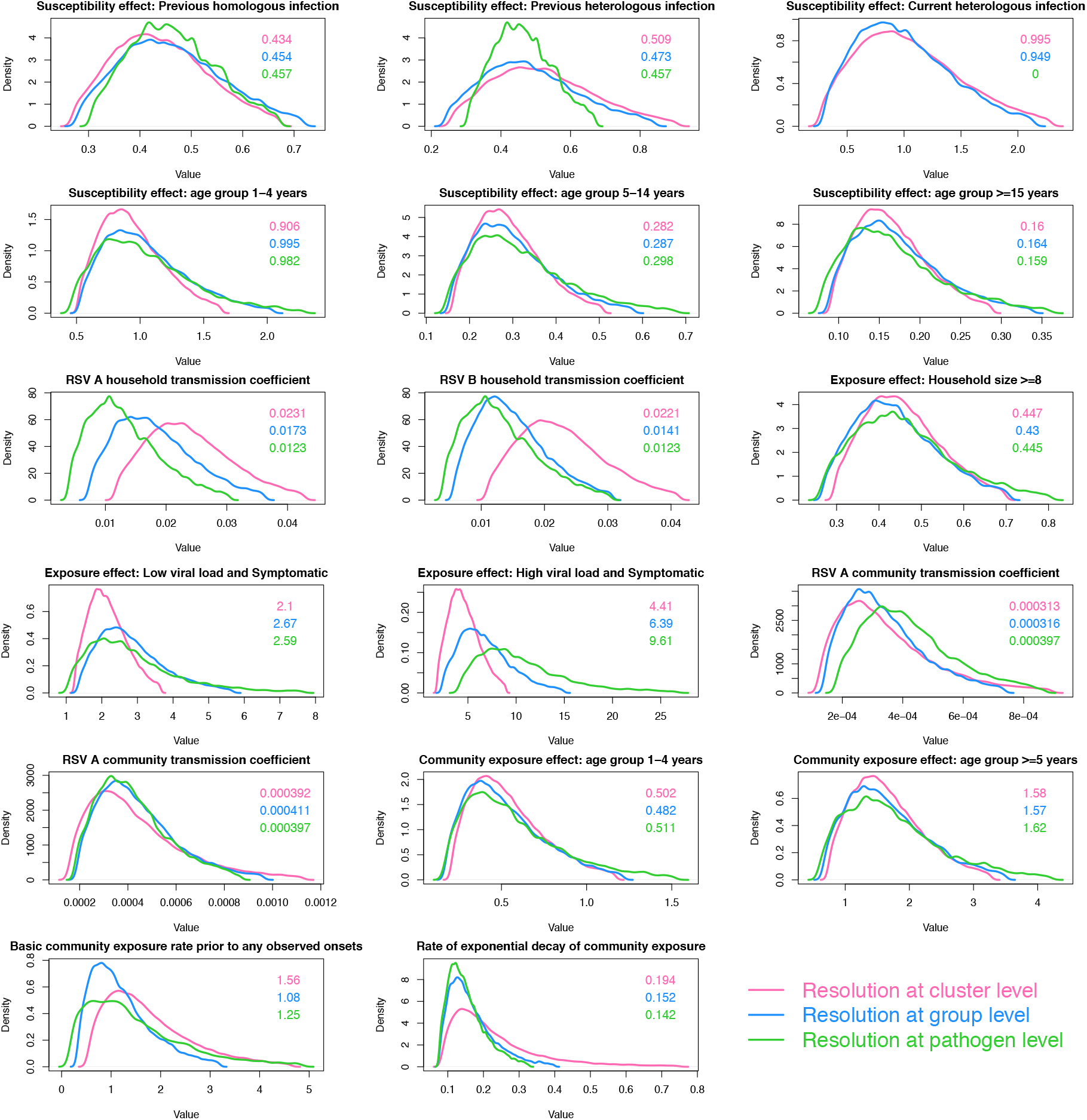
A comparison of the parameter distributions obtained from the model using different resolutions in pathogen identification. The green curves show the results using data at the pathogen level, the blue curves show the group level and the pink curves show the cluster level. Each panel shows 1 one of 17 comparable parameters. The values in the panel are the median parameter estimates colour-coded by pathogen resolution.

### Highest Probability transmission source

As described in the methods section, the HPTS was established for each case and these are shown in Figure 4. Thirty-nine out of ninety-seven (40%) of the RSV-A and 60/125 (48%) RSV-B cases were from sources outside of the household; 33% (13/39) of RSV-A introductions into the household led to infection of other household members, as did 38% (23/60) of RSV-B introductions. Table 1 gives the age distribution of all index cases compared to the age distribution of index cases that led to other infections in the household (HH outbreaks). Household outbreaks were as frequently initiated by a symptomatic infant as they were by a symptomatic child between 5-13 years. Fifty five percent (11/20) RSV-A and 36% (8/22) RSV-B infant (<1 year old) infections were acquired within the household. Of the 11 infant RSV-A, 8 were infected by children aged between 2 and 13 years (5 siblings and 3 cousins), 1 was infected by another younger infant (cousin), 1 by a 16-year old (unknown relation) and 1 by a 37-year old (mother). Five out of 8 of the infant RSV-B cases were infected by children between 2 and 13 years (4 siblings and 1 cousin), 2 were infected by a 16-year-old (unknown relation) and 18-year-old (sibling) while one was most likely infected by a 49-year-old (father). Figure 5 shows the transmission network by relationship centred around the infants. Infants infected several household members, mostly siblings and cousins.

**Table 1:**
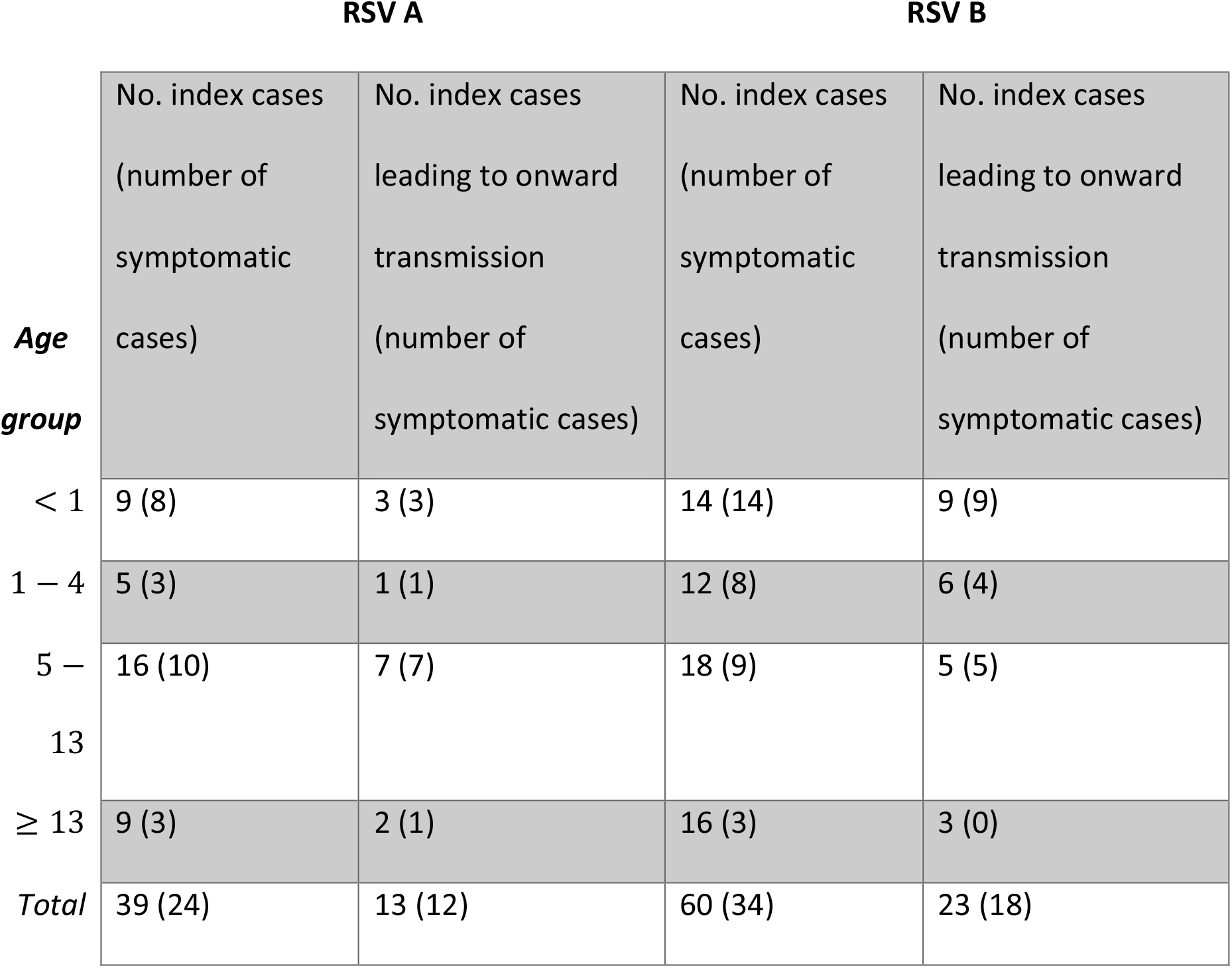
Age distribution of index cases of household outbreaks. Index cases are clustered into 4 age groups and according to whether they led to onward transmission in the household or not.

**Figure 4:**
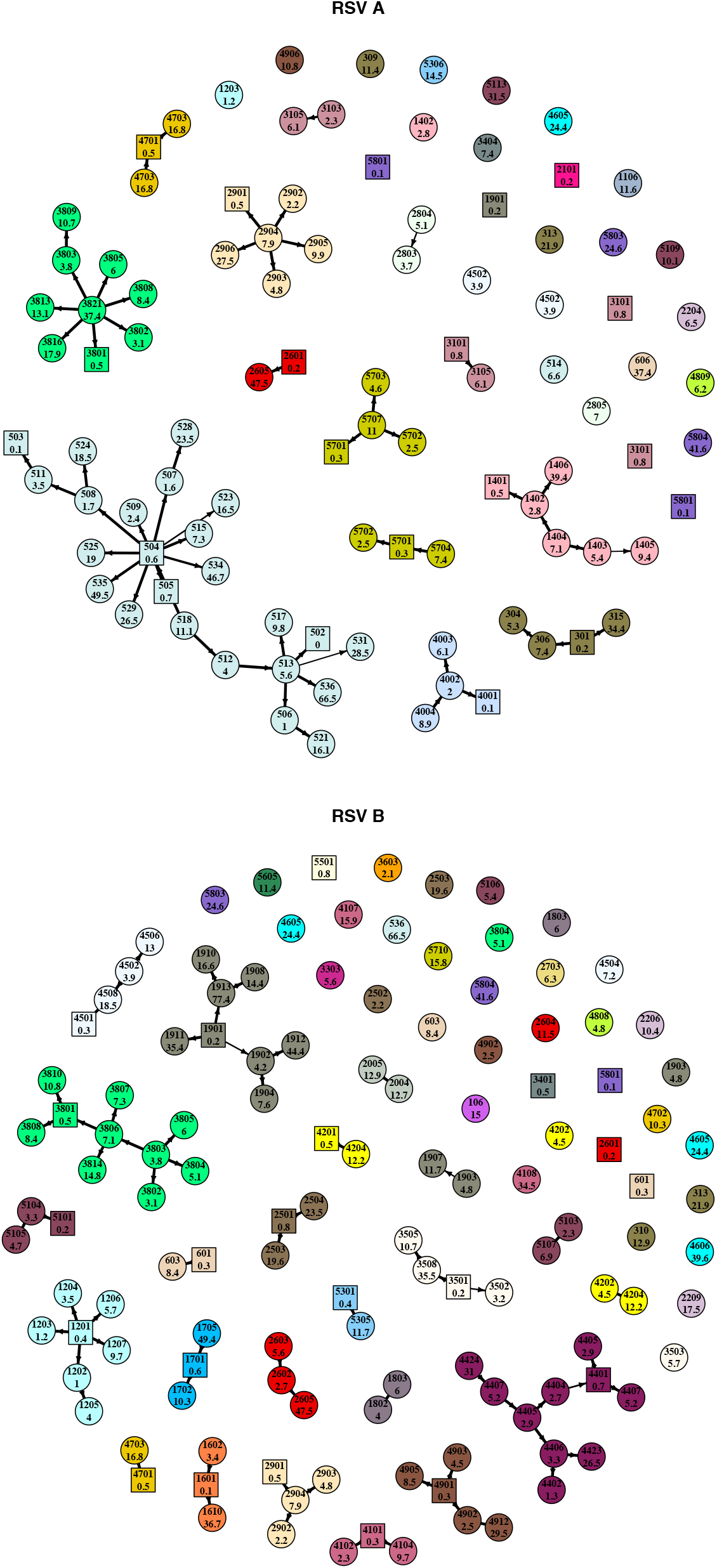
Transmission networks showing the highest probability source of transmission given by our model results. Each vertex is an RSV case labelled by individual study number (top) and age in years (bottom) and color-coded by household. Cases that are <1 year old are represented by square shaped vertices. The width of the connecting edge is proportional to the frequency at which the particular source was identified as the HPTS given different parameter set values.

**Figure 5:**
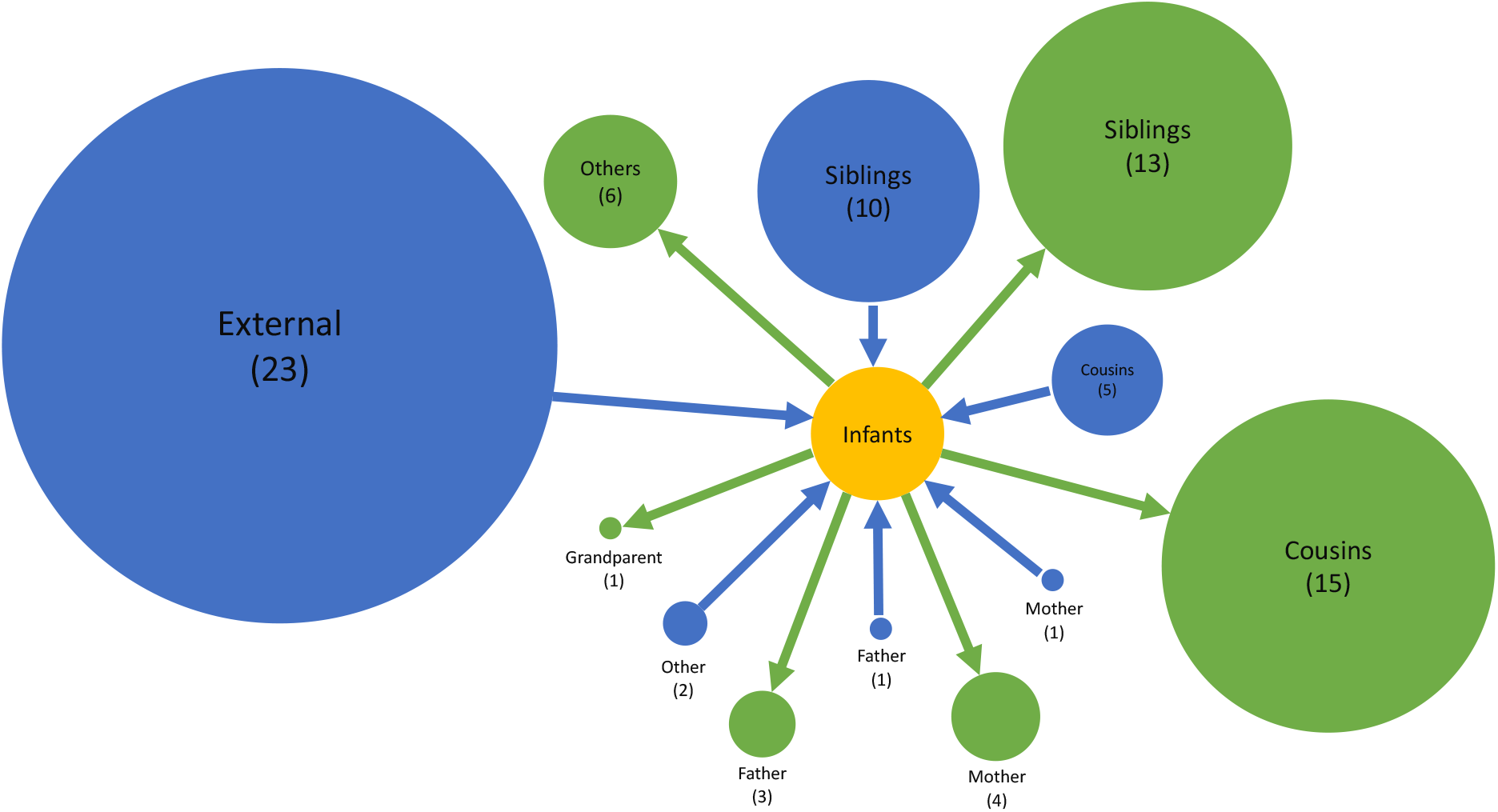
Network showing the sources of infection to the infant and who the infants infected as identified by social relationship. The blue circles show the sources to infant infection while the green show who the infants infected. The size of the circles is proportional to the number of cases which is given in brackets.

## Discussion

We carried out an analysis of data on the social-temporal and genetic pattern of spread of RSV in a sample of households in rural Kenya followed up for six months with an aim of identifying characteristics of household transmission chains. We found that most household outbreaks were initiated by a symptomatic child <13 years old, with infants and 5-13 year olds contributing equally. Infant infections that occurred in the household were mostly attributed to transmission from an elder sibling or cousin between 2 and 13 years old. Similar to a simulation study based on the same population from which our data was collected [19], we found over half of the infant infections were acquired outside of the household. Infants were the source of infection in 42/123 infections that were acquired in the household. Infants are therefore not only an important risk group but are also important as transmitters of household RSV infections. These results imply that a reduction in infant infections, say through a vaccine, would have a positive indirect (otherwise called herd) effect on RSV infections in other age groups. In addition, vaccination of household co-occupants of pre-school and school-going age would have an impact through reducing within household transmission to the infant. A significant portion of index cases that led to onward transmission in the household were symptomatic, a factor which we inferred to increase infectiousness. This implies - similar to past work [18]- that a vaccine that works against symptomatic infections, therefore reducing infectiousness, should be highly effective.

We inferred that 55% of infant RSV-A infections were acquired within the household, compared to 36% of infant RSV-B. There were also slight differences between the RSV groups in terms of proportion of cases that were index cases and proportion of index cases that led to onward transmission. Slower mutation rates in RSV-A [38,39], implying less variability from one season to the next, could account for its niche being in young infection-naïve infants as opposed to older individuals with previous exposure to RSV. In accord with this, White *et al* found evidence that RSV-A is slightly more transmissible than RSV-B [40].

We cannot state with certainty that there is a difference in transmission niche between the two groups, a study that incorporates information from different potential transmission hubs such as households, schools and workplaces would be better placed to do so. These differences between RSV-A and RSV-B might be specific to the outbreak under investigation, however, they do call for further investigations.

Through combining epidemiological and phylogenetic inference, our method was able to better resolve transmission chains within households compared to a preceding phylogenetic analysis [26,41]. The networks inferred from the present analysis did not contradict any of the inference from the phylogenetic analysis, with one exception. We assigned individual 3806 as the source of 3801’s RSV-B infection rather than 3805. In addition to considering the social grouping, infection window and genetic cluster, our approach also considers the infectiousness of a potential source. In this case, 3806 had symptoms and a high viral load in the three days preceding shedding onset in 3801, while 3805 did not. Such an example highlights the strength in our technique in being able to incorporate all possible determinants of a transmission event. It is worth mentioning that several super-spreader events were inferred. The model arrives at these networks based on the patterns in the available data. Though such events are plausible, to tease apart true super-spreader events from “convenience” networks, additional data on within household contacts would be needed, such as the kind collected by [42].

We found that increased pathogen resolution by including WGS data had a slight effect on both accuracy (resulting in narrower credible intervals for some parameters) and model inference (resulting in a change of transmission hypothesis). With resolution at the group level we had previously inferred possible niche separations between RSV-A and RSV-B based on overlapping but slightly different distributions of the transmission coefficients. Increased pathogen resolution resulted in a slight change in parameter distributions and this form of evidence was lost. Other inferred dynamics such as the effect of age, household size, previous infection remained relatively unchanged. The lack of a drastic effect of increased pathogen resolution could be due to the study design. The frequency in sampling, increased the accuracy of inferred onset dates, while, information on the social structuring of the population in the form of households provided information on some of the most frequent contacts each participant had. Since the genetic information’s clustering pattern mimicked the household structure, its utility was likely marginal. This result should not be surprising; [43] in integrating genetic, temporal and contact data found that contact data could replace the genetic data in transmission chain inference. This implies that good quality data on timing of cases and their most frequent contacts is key to be able to infer transmission characteristics. Nonetheless, it should be borne in mind that during an outbreak, it can be difficult to effectively gather contact data. In place of a dense sampling, integrating temporal and genetic data is the next best thing. Our results point to data integration being able to reduce measurement error (increase accuracy of parameter estimates) and provide information essential for correct inference (change interpretation of estimated parameters).

This study is not without its limitations. Firstly, similar to previous work [33,44,45], we used a two-step approach in our application of phylodynamics. This has the potential to lead to inconsistencies that would otherwise not occur with simultaneous inference of the evolutionary and epidemiological dynamics. However, given that we only used aggregated results of the phylogenetic analysis, in the form of clusters, and raw nucleotide distances as opposed to phylogenetic tree distances, we do not heavily rely on the exact results of the independent phylogenetic analysis. Using genetic clusters provides the advantage of being able to identify obvious separate introductions, a characteristic that can be difficult to account for in the models of simultaneous inference. The two-step approach was more computationally tractable than a simultaneous-inference version of it would have been. Secondly, the clusters were not probabilistically determined, in particular, uncertainty in the estimated date of sequence divergence was not considered. Finally, given the sampling interval of 3-4 days, short duration shedding episodes might have been missed and apparent co-index cases might actually have different onset dates.

In conclusion, we were able to integrate the results of a phylogenetic analysis with epidemiological data to infer that nearly half of the RSV infections in this study were acquired within the household. We showed explicitly that most infants were infected by an older sibling or cousin (2 to 13 years). A vaccine that limits the transmission capabilities (e.g. by eliminating ARI symptoms and reducing viral load) of this age group is therefore likely to reduce a significant portion of infant infection through indirect protection. The differences in infection patterns and interaction through modified susceptibility inferred between RSV-A and RSV-B warrant further investigation.

## Data Availability

The data used to generate the social-temporal shedding patterns is available under the Creative Commons Attribution 4.0 International (CC BY 4.0). For access to data and more detailed information beyond the metadata provided, there is a process of managed access requiring submission of a request form for consideration by the Data Governance Committee at KEMRI-Wellcome Trust Research Programme (http://kemri-wellcome.org/about-us/#ChildVerticalTab_15).
The sequence data used in this study can be found in both GenBank and Short Read Archive databases (see accession details in the Supplementary Dataset for the original manuscript https://www.nature.com/articles/s41598-019-46509-w).

http://kemri-wellcome.org/about-us/#ChildVerticalTab_15

https://www.nature.com/articles/s41598-019-46509-w

## Acknowledgements

We would like to thank Dr Anne Cori, Prof. Xavier Didelot and Dr George Githinji for their comments on the analysis. This paper is published with the permission of the director of Kenya Medical Research Institute.

